# Comparing SARS-CoV-2 natural immunity to vaccine-induced immunity: reinfections versus breakthrough infections

**DOI:** 10.1101/2021.08.24.21262415

**Authors:** Sivan Gazit, Roei Shlezinger, Galit Perez, Roni Lotan, Asaf Peretz, Amir Ben-Tov, Dani Cohen, Khitam Muhsen, Gabriel Chodick, Tal Patalon

## Abstract

**Background:** Reports of waning vaccine-induced immunity against COVID-19 have begun to surface. With that, the comparable long-term protection conferred by previous infection with SARS-CoV-2 remains unclear.

**Methods:** We conducted a retrospective observational study comparing three groups: (1)SARS-CoV-2-naïve individuals who received a two-dose regimen of the BioNTech/Pfizer mRNA BNT162b2 vaccine, (2)previously infected individuals who have not been vaccinated, and (3)previously infected *and* single dose vaccinated individuals. Three multivariate logistic regression models were applied. In all models we evaluated four outcomes: SARS-CoV-2 infection, symptomatic disease, COVID-19-related hospitalization and death. The follow-up period of June 1 to August 14, 2021, when the Delta variant was dominant in Israel.

**Results:** SARS-CoV-2-naïve vaccinees had a 13.06-fold (95% CI, 8.08 to 21.11) increased risk for breakthrough infection with the Delta variant compared to those previously infected, when the first event (infection or vaccination) occurred during January and February of 2021. The increased risk was significant (*P*<0.001) for symptomatic disease as well. When allowing the infection to occur at any time before vaccination (from March 2020 to February 2021), evidence of waning natural immunity was demonstrated, though SARS-CoV-2 naïve vaccinees had a 5.96-fold (95% CI, 4.85 to 7.33) increased risk for breakthrough infection and a 7.13-fold (95% CI, 5.51 to 9.21) increased risk for symptomatic disease. SARS-CoV-2-naïve vaccinees were also at a greater risk for COVID-19-related-hospitalizations compared to those that were previously infected.

**Conclusions:** This study demonstrated that natural immunity confers longer lasting and stronger protection against infection, symptomatic disease and hospitalization caused by the Delta variant of SARS-CoV-2, compared to the BNT162b2 two-dose vaccine-induced immunity. Individuals who were both previously infected with SARS-CoV-2 and given a single dose of the vaccine gained additional protection against the Delta variant.

## Introduction

The heavy toll that SARS-CoV-2 infection has been taking on global health and healthcare resources has created an urgent need to estimate which part of the population is protected against COVID-19 at a given time in order to set healthcare policies such as lockdowns and to assess the possibility of herd immunity.

To date, there is still no evidence-based, long-term correlate of protection^1^. This lack of correlate of protection has led to different approaches in terms of vaccine resource allocation, namely the need for vaccine administration in recovered patients, the need for booster shots in previously vaccinated individuals or the need to vaccinate low-risk populations, potentially previously exposed.

The short-term effectiveness of a two-dose regimen of the BioNTech/Pfizer BNT162b2 mRNA COVID-19 vaccine was demonstrated in clinical trials^2^ and in observational settings^3,4^. However, long term effectiveness across different variants is still unknown, though reports of waning immunity are beginning to surface, not merely in terms of antibody dynamics over time^5–7^, but in real-world settings as well^8^. Alongside the question of long-term protection provided by the vaccine, the degree and duration to which previous infection with SARS-CoV-2 affords protection against repeated infection also remains unclear. Apart from the paucity of studies examining long-term protection against reinfection^9^, there is a challenge in defining reinfection as opposed to prolonged viral shedding^10^. While clear-cut cases exist, namely two separate clinical events with two distinct sequenced viruses, relying solely on these cases will likely result in an under-estimation of the incidence of reinfection.

Different criteria based on more widely-available information have been suggested^11^, the Centers for Disease Control and Prevention’s (CDC) guidelines refer to two positive SARS-CoV-2 polymerase chain reaction (PCR) test results at least 90 days apart.^12^ Using similar criteria, population-based studies demonstrated natural immunity^13,14^ with no signs of waning immunity for at least 7 months, though protection was lower for those aged 65 or older^9^.

The Delta (B.1.617.2) Variant of Concern (VOC), initially identified in India and today globally prevalent, has been the dominant strain in Israel since June 2021. The recent surge of cases in Israel^15^, one of the first countries to embark on a nationwide vaccination campaign (mostly with the BioNTech/Pfizer BNT162b2 vaccine), has raised concerns about vaccine effectiveness against the Delta variant, including official reports of decreased protection^16^. Concomitantly, studies have demonstrated only mild differences in short-term vaccine effectiveness^17^ against the Delta variant, as well as substantial antibody response^18^. Apart from the variant, the new surge was also explained by the correlation found between time-from-vaccine and breakthrough infection rates, as early vaccinees were demonstrated to be significantly more at risk than late vaccinees^8^. Now, when sufficient time has passed since both the beginning of the pandemic and the deployment of the vaccine, we can examine the long-term protection of natural immunity compared to vaccine-induced immunity.

To this end, we compared the incidence rates of breakthrough infections to the incidence rates of reinfection, leveraging the centralized computerized database of Maccabi Healthcare Services (MHS), Israel’s second largest Health Maintenance Organization.

## Methods

### Study design and population

A retrospective cohort study was conducted, leveraging data from MHS’ centralized computerized database. The study population included MHS members aged 16 or older who were vaccinated prior to February 28, 2021, who had a documented SARS-CoV-2 infection by February 28, 2021, or who had both a documented SARS-CoV-2 infection by February 28, 2021 *and* received one dose of the vaccine by May 25, 2021, at least 7 days before the study period. On March 2, 2021, The Israeli Ministry of Health revised its guidelines and allowed previously SARS-CoV-2 infected individuals to receive one dose of the vaccine, after a minimum 3-month-interval from the date of infection

### Data Sources

Anonymized Electronic Medical Records (EMRs) were retrieved from MHS’ centralized computerized database for the study period of March 1, 2020 to August 14, 2021.

MHS is a 2.5-million-member, state-mandated, non-for-profit, second largest health fund in Israel, which covers 26% of the population and provides a representative sample of the Israeli population. Membership in one of the four national health funds is mandatory, whereas all citizens must freely choose one of four funds, which are prohibited by law from denying membership to any resident. MHS has maintained a centralized database of EMRs for three decades, with less than 1% disengagement rate among its members, allowing for a comprehensive longitudinal medical follow-up. The centralized dataset includes extensive demographic data, clinical measurements, outpatient and hospital diagnoses and procedures, medications dispensed, imaging performed and comprehensive laboratory data from a single central laboratory.

### Data extraction and definition of the study variables

#### COVID-19-related data

COVID-19-related information was captured as well, including dates of the first and second dose of the vaccine and results of any polymerase chain reaction (PCR) tests for SARS-CoV-2, given that all such tests are recorded centrally. Records of COVID-19-related hospitalizations were retrieved as well, and COVID-19-related mortality was screened for. Additionally, information about COVID-19-related symptoms was extracted from EMRs, where they were recorded by the primary care physician or a certified nurse who conducted in-person or phone visits with each infected individual.

#### Exposure variable: study groups

The eligible study population was divided into three groups: (1)fully vaccinated and SARS-CoV-2-naïve individuals, namely MHS members who received two doses of the BioNTech/Pfizer mRNA BNT162b2 vaccine by February 28, 2021, did not receive the third dose by the end of the study period and did not have a positive PCR test result by June 1, 2021; (2) unvaccinated previously infected individuals, namely MHS members who had a positive SARS-CoV-2 PCR test recorded by February 28, 2021 and who had not been vaccinated by the end of the study period; (3) previously infected *and* vaccinated individuals, including individuals who had a positive SARS-CoV-2 PCR test by February 28, 2021 and received one dose of the vaccine by May 25, 2021, at least 7 days before the study period. The fully vaccinated group was the comparison (reference) group in our study. Groups 2 and 3, were matched to the comparison group 1 in a 1:1 ratio based on age, sex and residential socioeconomic status.

#### Dependent variables

We evaluated four SARS-CoV-2-related outcomes, or second events: documented RT-PCR confirmed SARS-CoV-2 infection, COVID-19, COVID-19-related hospitalization and death. Outcomes were evaluated during the follow-up period of June 1 to August 14, 2021, the date of analysis, corresponding to the time in which the Delta variant became dominant in Israel.

#### Covariates

Individual-level data of the study population included patient demographics, namely age, sex, socioeconomic status (SES) and a coded geographical statistical area (GSA, assigned by Israel’s National Bureau of Statistics, corresponds to neighborhoods and is the smallest geostatistical unit of the Israeli census). The SES is measured on a scale from 1 (lowest) to 10, and the index is based on several parameters, including household income, educational qualifications, household crowding and car ownership. Data were also collected on last documented body mass index (BMI) and information about chronic diseases from MHS’ automated registries, including cardiovascular diseases^19^, hypertension^20^, diabetes^21^, chronic kidney disease^22^, chronic obstructive pulmonary disease, immunocompromised conditions, and cancer from the National Cancer Registry^23^.

### Statistical analysis

Two multivariate logistic regression models were applied that evaluated the four aforementioned SARS-CoV-2-related outcomes as dependent variables, while the study groups were the main independent variables.

#### Model 1– previously infected vs. vaccinated individuals, with matching for time of first event

In model 1, we examined natural immunity and vaccine-induced immunity by comparing the likelihood of SARS-CoV-2-related outcomes between previously infected individuals who have never been vaccinated and fully vaccinated SARS-CoV-2-naïve individuals. These groups were matched in a 1:1 ratio by age, sex, GSA and time of first event. The first event (the preliminary exposure) was either the time of administration of the second dose of the vaccine *or* the time of documented infection with SARS-CoV-2 (a positive RT-PCR test result), both occurring between January 1, 2021 and February 28, 2021. Thereby, we matched the “immune activation” time of both groups, examining the long-term protection conferred when vaccination or infection occurred within the same time period. The three-month interval between the first event and the second event was implemented in order to capture reinfections (as opposed to prolonged viral shedding) by following the 90-day guideline of the CDC.

#### Model 2

In model 2, we compared the SARS-CoV-2 naïve vaccinees to unvaccinated previously infected individuals while intentionally *not* matching the time of the first event (i.e., either vaccination or infection), in order to compare vaccine-induced immunity to natural immunity, regardless of time of infection. Therefore, matching was done in a 1:1 ratio based on age, sex and GSA alone. Similar to the model 1, either event (vaccination or infection) had to occur by February 28, to allow for the 90-day interval. The four SARS-CoV-2 study outcomes were the same for this model, evaluated during the same follow-up period.

#### Model 3

Model 3 examined previously infected individuals vs. previously-infected-and-once-vaccinated individuals, using “natural immunity” as the baseline group. We matched the groups in a 1:1 ratio based on age, sex and GSA. SARS-CoV-2 outcomes were the same, evaluated during the same follow-up period.

In all three models, we estimated natural immunity vs. vaccine-induced immunity for each SARS-CoV-2-related outcome, by applying logistic regression to calculate the odds ratio (OR) between the two groups in each model, with associated 95% confidence intervals (CIs). Results were then adjusted for underlying comorbidities, including obesity, cardiovascular diseases, diabetes, hypertension, chronic kidney disease, cancer and immunosuppression conditions.

Analyses were performed using Python version 3.73 with the stats model package. *P*□<□0.05 was considered statistically significant.

#### Ethics declaration

This study was approved by the MHS (Maccabi Healthcare Services) Institutional Review Board (IRB). Due to the retrospective design of the study, informed consent was waived by the IRB, and all identifying details of the participants were removed before computational analysis.

#### Data availability statement

According to the Israel Ministry of Health regulations, individual-level data cannot be shared openly. Specific requests for remote access to de-identified community-level data should be directed to KSM, Maccabi Healthcare Services Research and Innovation Center.

#### Code availability

Specific requests for remote access to the code used for data analysis should be referred to KSM, Maccabi Healthcare Services Research and Innovation Center.

## Results

Overall, 673,676 MHS members 16 years and older were eligible for the study group of fully vaccinated SARS-CoV-2-naïve individuals; 62,883 were eligible for the study group of unvaccinated previously infected individuals and 42,099 individuals were eligible for the study group of previously infected and single-dose vaccinees.

### Model 1 – previously infected vs. vaccinated individuals, with matching for time of first event

In model 1, we matched 16,215 persons in each group. Overall, demographic characteristics were similar between the groups, with some differences in their comorbidity profile (Table 1a).

**Table 1a.** Characteristics of study population, model 1 and 2.

|  | **Model 1 – with matching of time of first event** |  | **Model 2 – without matching of time of first event** |  |
| --- | --- | --- | --- | --- |
| Characteristics | Previously infected<br>(n=16,215) | Vaccinated individuals<br>(n=16,215) | Previously infected<br>(n=46,035) | Previously infected *and* vaccinated<br>(n =46,035) |
| **Age years, mean (SD)** | 36.1 (13.9) | 36.1 (13.9) | 36.1 (14.7) | 36.1 (14.7) |
| **Age group – no. (%)** |  |  |  |  |
| 16 to 39 yr | 9,889 (61.0) | 9,889 (61.0) | 28,157 (61.2) | 28,157 (61.2) |
| 40 to 59 yr | 5,536 (34.1) | 5,536 (34.1) | 14,973 (32.5) | 14,973 (32.5) |
| ≥60 yr | 790 (4.9) | 790 (4.9) | 2,905 (6.3) | 2,905 (6.3) |
| **Sex – no. (%)** |  |  |  |  |
| Female | 7,428 (45.8) | 7,428 (45.8) | 22,661 (49.2) | 22,661 (49.2) |
| Male | 8,787 (54.2) | 8,787 (54.2) | 23,374 (50.8) | 23,374 (50.8) |
| **SES, mean (SD)** | 5.5 (1.9) | 5.5 (1.9) | 5.3 (1.9) | 5.3 (1.9) |
| **Comorbidities – no. (%)** |  |  |  |  |
| Hypertension | 1,276 (7.9) | 1,569 (9.7) | 4,009 (8.7) | 4,301 (9.3) |
| CVD | 551 (3.4) | 647 (4.0) | 1,875 (4.1) | 1830 (4.0) |
| DM | 635 (3.9) | 877 (5.4) | 2207 (4.8) | 2300 (5.0) |
| Immunocompromised | 164 (1.0) | 420 (2.6) | 527 (1.1) | 849 (1.8) |
| Obesity (BMI ≥30) | 3,076 (19.0) | 3,073 (19.0) | 9,117 (19.8) | 8,610 (18.7) |
| CKD | 196 (1.2) | 271 (1.7) | 659 (1.4) | 814 (1.8) |
| COPD | 65 (0.4) | 97 (0.6) | 218 (0.5) | 292 (0.6) |
| Cancer | 324 (2.0) | 636 (3.9) | 1,044 (2.3) | 1,364 (3.0) |

SD – Standard Deviation; SES – Socioeconomic status on a scale from 1 (lowest) to 10; CVD – Cardiovascular Diseases; DM – Diabetes Mellitus; CKD – Chronic Kidney Disease; COPD – Chronic Obstructive Pulmonary Disease.

**Table 1b.**
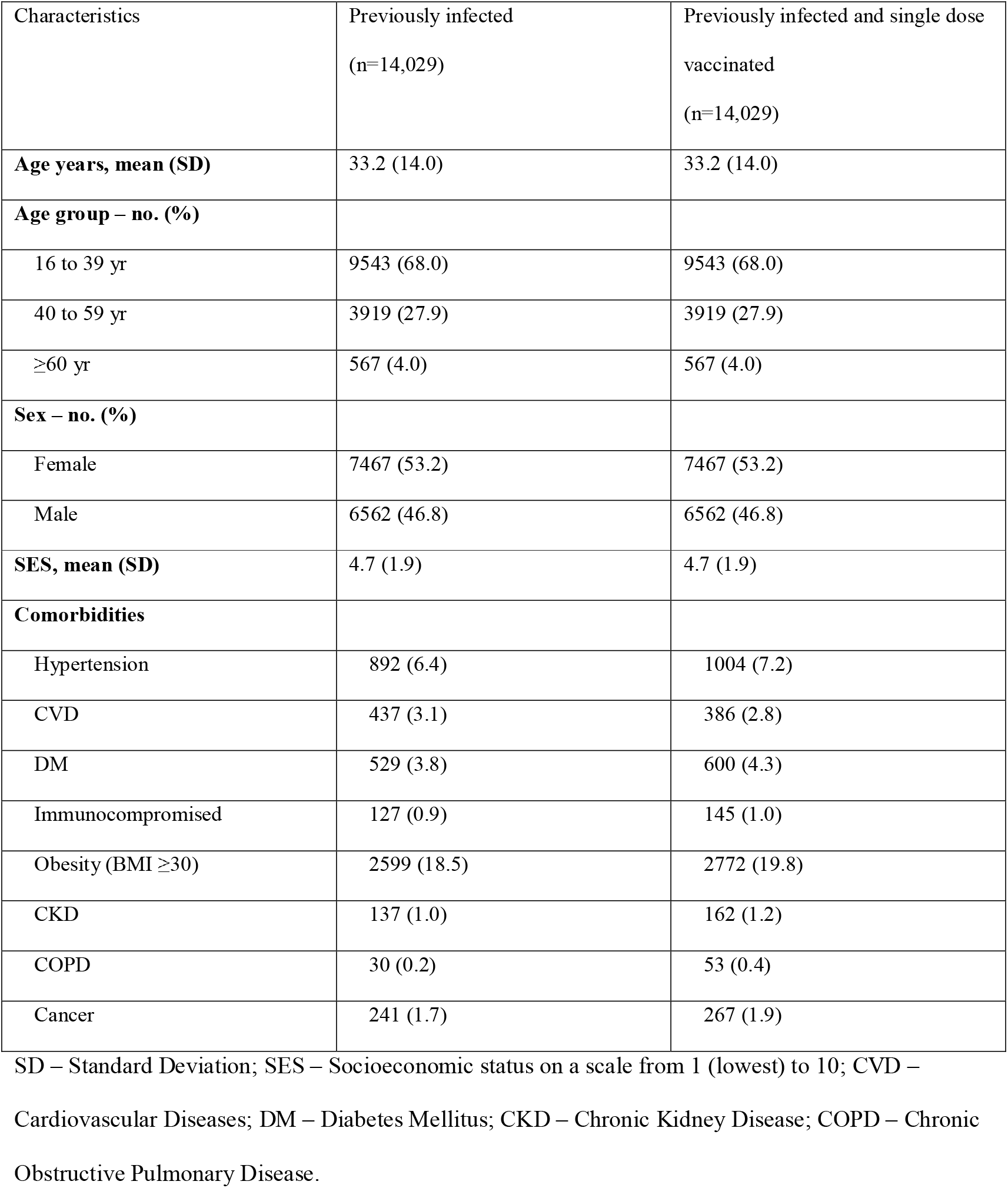
Characteristics of study population, model 3.

| Characteristics | Previously infected<br>(n=14,029) | Previously infected and single dose<br>vaccinated<br>(n=14,029) |
| --- | --- | --- |
| **Age years, mean (SD)** | 33.2 (14.0) | 33.2 (14.0) |
| **Age group – no. (%)** |  |  |
| 16 to 39 yr | 9543 (68.0) | 9543 (68.0) |
| 40 to 59 yr | 3919 (27.9) | 3919 (27.9) |
| ≥60 yr | 567 (4.0) | 567 (4.0) |
| **Sex – no. (%)** |  |  |
| Female | 7467 (53.2) | 7467 (53.2) |
| Male | 6562 (46.8) | 6562 (46.8) |
| **SES, mean (SD)** | 4.7 (1.9) | 4.7 (1.9) |
| **Comorbidities** |  |  |
| Hypertension | 892 (6.4) | 1004 (7.2) |
| CVD | 437 (3.1) | 386 (2.8) |
| DM | 529 (3.8) | 600 (4.3) |
| Immunocompromised | 127 (0.9) | 145 (1.0) |
| Obesity (BMI ≥30) | 2599 (18.5) | 2772 (19.8) |
| CKD | 137 (1.0) | 162 (1.2) |
| COPD | 30 (0.2) | 53 (0.4) |
| Cancer | 241 (1.7) | 267 (1.9) |

SD – Standard Deviation; SES – Socioeconomic status on a scale from 1 (lowest) to 10; CVD –
Cardiovascular Diseases; DM – Diabetes Mellitus; CKD – Chronic Kidney Disease; COPD – Chronic Obstructive Pulmonary Disease.

During the follow-up period, 257 cases of SARS-CoV-2 infection were recorded, of which 238 occurred in the vaccinated group (breakthrough infections) and 19 in the previously infected group (reinfections). After adjusting for comorbidities, we found a statistically significant 13.06-fold (95% CI, 8.08 to 21.11) increased risk for breakthrough infection as opposed to reinfection (*P*<0.001). Apart from age ≥60 years, there was no statistical evidence that any of the assessed comorbidities significantly affected the risk of an infection during the follow-up period (Table 2a). As for symptomatic SARS-COV-2 infections during the follow-up period, 199 cases were recorded, 191 of which were in the vaccinated group and 8 in the previously infected group. Symptoms for all analyses were recorded in the central database within 5 days of the positive RT-PCR test for 90% of the patients, and included chiefly fever, cough, breathing difficulties, diarrhea, loss of taste or smell, myalgia, weakness, headache and sore throat. After adjusting for comorbidities, we found a 27.02-fold risk (95% CI, 12.7 to 57.5) for symptomatic breakthrough infection as opposed to symptomatic reinfection (*P*<0.001) (Table 2b). None of the covariates were significant, except for age ≥60 years.

**Table 2a.**
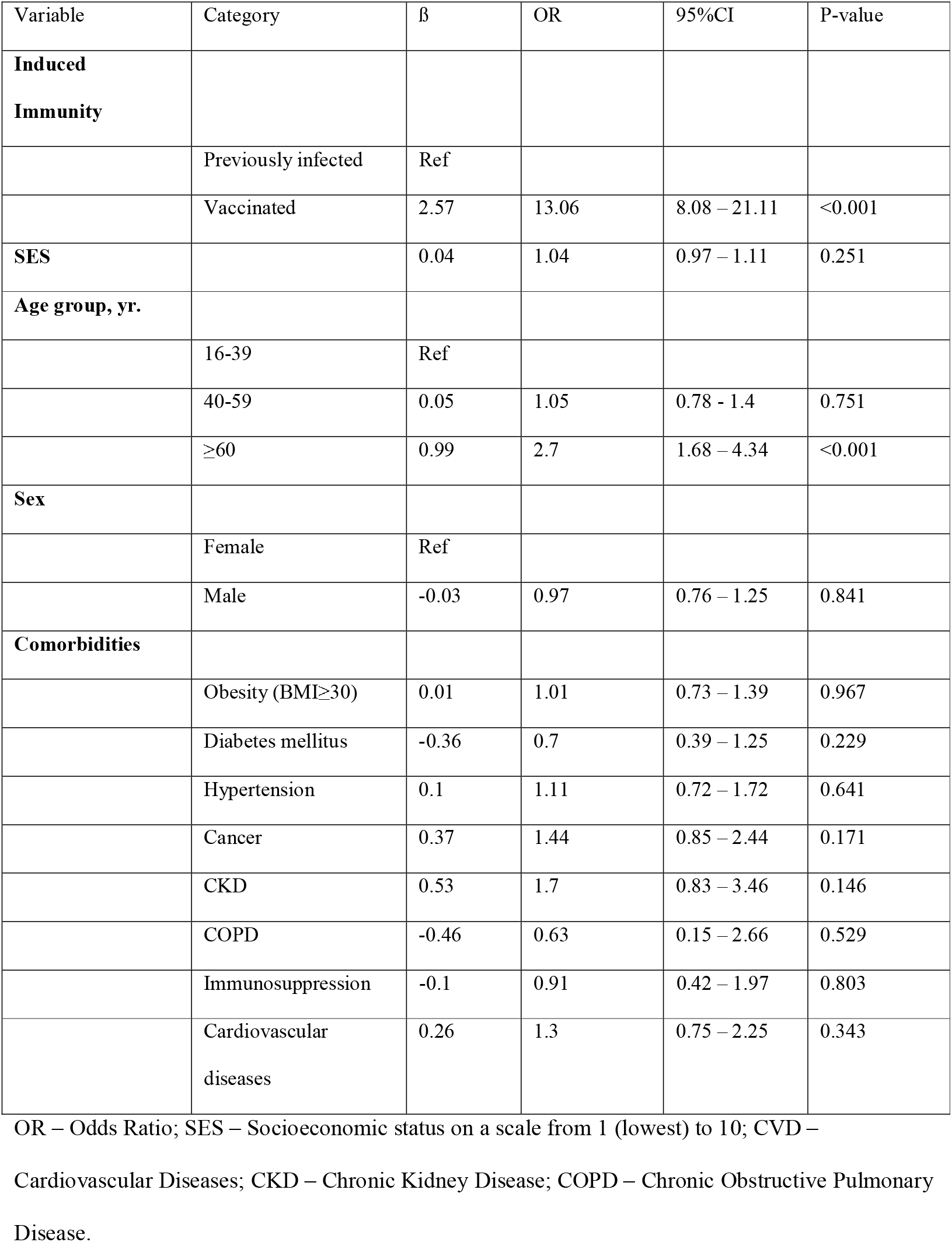
OR for SARS-CoV-2 infection, model 1, previously infected vs. vaccinated

**Table 2b.** OR for Symptomatic SARS-CoV-2 infection, model 1, previously infected vs. vaccinated

| Variable | Category | $\beta$ | OR | 95%CI | P-value |
| --- | --- | --- | --- | --- | --- |
| **Induced Immunity** |  |  |  |  |  |
|  | Previously infected | Ref |  |  |  |
|  | Vaccinated | 3.3 | 27.02 | 12.7 – 57.5 | <0.001 |
| **SES** |  | 0.04 | 1.04 | 0.96 – 1.12 | 0.312 |
| **Age group, yr.** |  |  |  |  |  |
|  | 16-39 | Ref |  |  |  |
|  | 40-59 | 0.19 | 1.21 | 0.88 – 1.67 | 0.25 |
|  | ≥60 | 1.06 | 2.89 | 1.68 – 4.99 | <0.001 |
| **Sex** |  |  |  |  |  |
|  | Female | Ref |  |  |  |
|  | Male | -0.19 | 0.82 | 0.62 – 1.1 | 0.185 |
| **Comorbidities** |  |  |  |  |  |
|  | Obesity (BMI≥30) | 0.02 | 1.02 | 0.71 – 1.48 | 0.899 |
|  | Diabetes mellitus | -0.31 | 0.73 | 0.37 – 1.43 | 0.361 |
|  | Hypertension | 0.12 | 1.13 | 0.69 – 1.85 | 0.623 |
|  | Cancer | 0.37 | 1.45 | 0.8 – 2.62 | 0.217 |
|  | CKD | 0.1 | 1.1 | 0.42 – 2.87 | 0.846 |
|  | COPD | -0.78 | 0.46 | 0.06 – 3.41 | 0.445 |
|  | Immunosuppression | -0.37 | 0.69 | 0.25 – 1.89 | 0.468 |
|  | Cardiovascular diseases | 0.03 | 1.03 | 0.52 – 2.03 | 0.941 |

OR – Odds Ratio; SES – Socioeconomic status on a scale from 1 (lowest) to 10; CVD –
Cardiovascular Diseases; CKD – Chronic Kidney Disease; COPD – Chronic Obstructive Pulmonary Disease.

Nine cases of COVID-19-related hospitalizations were recorded, 8 of which were in the vaccinated group and 1 in the previously infected group (Table S1). No COVID-19-related deaths were recorded in our cohorts.

### Model 2 –previously infected vs. vaccinated individuals, without matching for time of first event

In model 2, we matched 46,035 persons in each of the groups (previously infected vs. vaccinated). Baseline characteristics of the groups are presented in Table 1a. Figure 1 demonstrates the timely distribution of the first infection in reinfected individuals. When comparing the vaccinated individuals to those previously infected at any time (including during 2020), we found that throughout the follow-up period, 748 cases of SARS-CoV-2 infection were recorded, 640 of which were in the vaccinated group (breakthrough infections) and 108 in the previously infected group (reinfections). After adjusting for comorbidities, a 5.96-fold increased risk (95% CI, 4.85 to 7.33) increased risk for breakthrough infection as opposed to reinfection could be observed (*P*<0.001) (Table 3a). Apart from SES level and age ≥60, that remained significant in this model as well, there was no statistical evidence that any of the comorbidities significantly affected the risk of an infection.

**Table 3a.** OR for SARS-CoV-2 infection, model 2, previously infected vs. vaccinated

| Variable | Category | $\beta$ | OR | 95%CI | P-value |
| --- | --- | --- | --- | --- | --- |
| **Induced Immunity** |  |  |  |  |  |
|  | Previously infected | Ref |  |  |  |
|  | Vaccinated | 1.78 | 5.96 | 4.85 – 7.33 | <0.001 |
| **SES** |  | 0.07 | 1.07 | 1.03 – 1.11 | <0.001 |
| **Age group, yr.** |  |  |  |  |  |
|  | 16-39 | Ref |  |  |  |
|  | 40-59 | 0.06 | 1.06 | 0.9 – 1.26 | 0.481 |
| | $\geq 60$ | 0.79 | 2.2 | 1.66 – 2.92 | <0.001 |
| **Sex** |  |  |  |  |  |
|  | Female | Ref |  |  |  |
|  | Male | -0.01 | 0.99 | 0.85 - 1.14 | 0.842 |
| **Comorbidities** |  |  |  |  |  |
| | Obesity (BMI$\geq 30$) | 0.12 | 1.13 | 0.94 – 1.36 | 0.202 |
|  | Diabetes mellitus | -0.15 | 0.86 | 0.61 – 1.22 | 0.4 |
|  | Hypertension | -0.12 | 0.89 | 0.67 – 1.17 | 0.402 |
|  | Cancer | 0.2 | 1.22 | 0.85 – 1.76 | 0.283 |
|  | CKD | 0.3 | 1.35 | 0.85 – 2.14 | 0.207 |
|  | COPD | 0.48 | 1.62 | 0.88 – 2.97 | 0.121 |
|  | Immunosuppression | -0.03 | 0.98 | 0.57 – 1.66 | 0.925 |
|  | Cardiovascular diseases | 0.08 | 1.09 | 0.77 – 1.53 | 0.638 |

OR – Odds Ratio; SES – Socioeconomic status on a scale from 1 (lowest) to 10; CVD –
Cardiovascular Diseases; CKD – Chronic Kidney Disease; COPD – Chronic Obstructive Pulmonary Disease.

**Figure 1.**
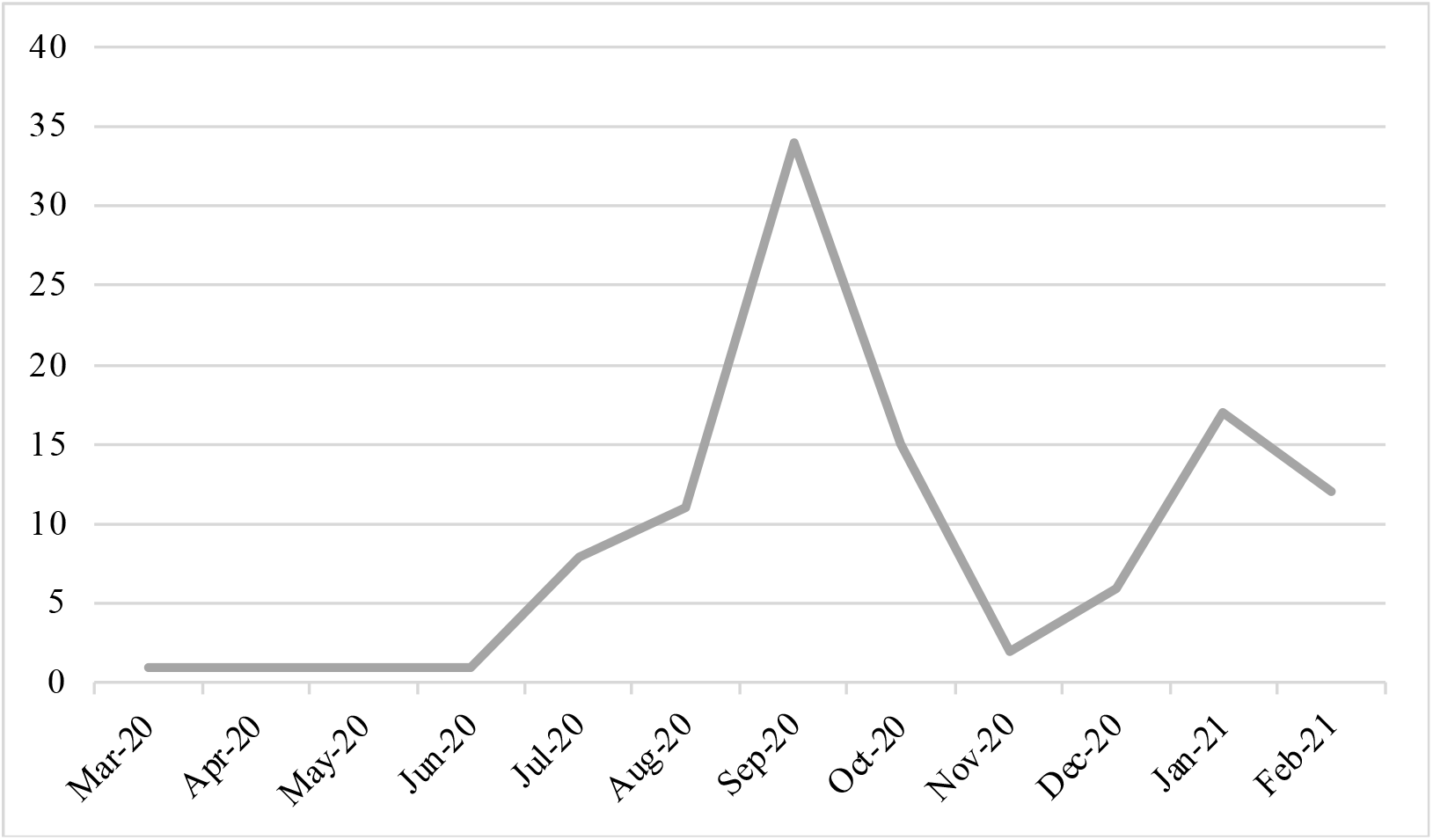
Time of first infection in those reinfected between June and August 2021, model 2.

Overall, 552 symptomatic cases of SARS-CoV-2 were recorded, 484 in the vaccinated group and 68 in the previously infected group. There was a 7.13-fold (95% CI, 5.51 to 9.21) increased risk for symptomatic breakthrough infection than symptomatic reinfection (Table 3b). COVID-19 related hospitalizations occurred in 4 and 21 of the reinfection and breakthrough infection groups, respectively. Vaccinated individuals had a 6.7-fold (95% CI, 1.99 to 22.56) increased to be admitted compared to recovered individuals. Being 60 years of age or older significantly increased the risk of COVID-19-related hospitalizations (Table S2). No COVID-19-related deaths were recorded.

**Table 3b.** OR for Symptomatic SARS-CoV-2 infection, model 2, previously infected vs. vaccinated

| Variable | Category | $\beta$ | OR | 95%CI | P-value |
| --- | --- | --- | --- | --- | --- |
| **Induced Immunity** |  |  |  |  |  |
|  | Previously infected | Ref |  |  |  |
|  | Vaccinated | 1.96 | 7.13 | 5.51 – 9.21 | <0.001 |
| **SES** |  | 0.07 | 1.07 | 1.02 – 1.12 | 0.003 |
| **Age group, yr.** |  |  |  |  |  |
|  | 16-39 | Ref |  |  |  |
|  | 40-59 | 0.09 | 1.1 | 0.9 – 1.33 | 0.35 |
|  | ≥60 | 0.8 | 2.23 | 1.61 – 3.09 | <0.001 |
| **Sex** |  |  |  |  |  |
|  | Female | Ref |  |  |  |
|  | Male | -0.02 | 0.98 | 0.82 – 1.16 | 0.785 |
| **Comorbidities** |  |  |  |  |  |
|  | Obesity (BMI≥30) | 0.16 | 1.18 | 0.95 – 1.46 | 0.133 |
|  | Diabetes mellitus | -0.11 | 0.89 | 0.61 – 1.32 | 0.571 |
|  | Hypertension | -0.01 | 0.99 | 0.72 – 1.35 | 0.943 |
|  | Cancer | 0.08 | 1.09 | 0.7 – 1.69 | 0.71 |
|  | CKD | 0.13 | 1.14 | 0.65 – 1.98 | 0.654 |
|  | COPD | 0.5 | 1.65 | 0.82 – 3.31 | 0.162 |
|  | Immunosuppression | 0 | 1 | 0.54 – 1.85 | 0.999 |
|  | Cardiovascular diseases | 0 | 1 | 0.67 – 1.5 | 0.99 |

OR – Odds Ratio; SES – Socioeconomic status on a scale from 1 (lowest) to 10; CVD –
Cardiovascular Diseases; CKD – Chronic Kidney Disease; COPD – Chronic Obstructive Pulmonary Disease.

### Model 3 - previously infected vs. vaccinated and previously infected individuals

In model 3, we matched 14,029 persons. Baseline characteristics of the groups are presented in Table 1b. Examining previously infected individuals to those who were both previously infected and received a single dose of the vaccine, we found that the latter group had a significant 0.53-fold (95% CI, 0.3 to 0.92) (Table 4a) decreased risk for reinfection, as 20 had a positive RT-PCR test, compared to 37 in the previously infected and unvaccinated group. Symptomatic disease was present in 16 single dose vaccinees and in 23 of their unvaccinated counterparts. One COVID-19-related hospitalization occurred in the unvaccinated previously infected group. No COVID-19-related mortality was recorded.

**Table 4a.** OR for SARS-CoV-2 infection, model 3, previously infected vs. previously infected and single-dose-vaccinated

| Variable | Category | $\beta$ | OR | 95%CI | P-value |
| --- | --- | --- | --- | --- | --- |
| **Induced Immunity** |  |  |  |  |  |
|  | Previously infected | Ref |  |  |  |
|  | Previously infected and vaccinated | -0.64 | 0.53 | 0.3 – 0.92 | 0.024 |
| **SES** |  | 0.11 | 1.12 | 0.98 – 1.28 | 0.096 |
| **Age group, yr.** |  |  |  |  |  |
|  | 16-59 | Ref |  |  |  |
| | $\geq 60$ | -0.81 | 0.44 | 0.06 – 3.22 | 0.422 |
| **Comorbidities** |  |  |  |  |  |
|  | Immunosuppression | 0.72 | 2.06 | 0.28 – 15.01 | 0.475 |

SES – Socioeconomic status on a scale from 1 (lowest) to 10

**Table 4b.** OR for Symptomatic SARS-CoV-2 infection, model 2, previously infected vs. previously infected and vaccinated

| Variable | Category | $\beta$ | OR | 95%CI | P-value |
| --- | --- | --- | --- | --- | --- |
| **Induced Immunity** |  |  |  |  |  |
|  | Previously infected | Ref |  |  |  |
|  | Previously infected and vaccinated | -0.43 | 0.65 | 0.34 – 1.25 | 0.194 |
| **SES** |  | 0.06 | 1.06 | 0.9 – 1.24 | 0.508 |
| **Age group, yr.** |  |  |  |  |  |
|  | 16-59 | Ref |  |  |  |
| | $\geq 60$ | -16.9 | 0 | 0.0 – inf | 0.996 |
| **Comorbidities** |  |  |  |  |  |
|  | Immunosuppression | 1.15 | 3.14 | 0.43 – 23.01 | 0.26 |

OR – Odds Ratio; SES – Socioeconomic status on a scale from 1 (lowest) to 10.

We conducted a further sub-analysis, compelling the single-dose vaccine to be administered *after* the positive RT-PCR test. This subset represented 81% of the previously-infected-and-vaccinated study group. When performing this analysis, we found a similar, though not significant, trend of decreased risk of reinfection, with an OR of 0.68 (95% CI, 0.38 to 1.21, *P-*value=0.188).

## Discussion

This is the largest real-world observational study comparing natural immunity, gained through previous SARS-CoV-2 infection, to vaccine-induced immunity, afforded by the BNT162b2 mRNA vaccine. Our large cohort, enabled by Israel’s rapid rollout of the mass-vaccination campaign, allowed us to investigate the risk for additional infection – either a breakthrough infection in vaccinated individuals or reinfection in previously infected ones – over a longer period than thus far described.

Our analysis demonstrates that SARS-CoV-2-naïve vaccinees had a 13.06-fold increased risk for breakthrough infection with the Delta variant compared to those previously infected, when the first event (infection or vaccination) occurred during January and February of 2021. The increased risk was significant for a symptomatic disease as well.

Broadening the research question to examine the extent of the phenomenon, we allowed the infection to occur at any time between March 2020 to February 2021 (when different variants were dominant in Israel), compared to vaccination only in January and February 2021. Although the results could suggest waning natural immunity against the Delta variant, those vaccinated are still at a 5.96-fold increased risk for breakthrough infection and at a 7.13-fold increased risk for symptomatic disease compared to those previously infected. SARS-CoV-2-naïve vaccinees were also at a greater risk for COVID-19-related-hospitalization compared to those who were previously infected.

Individuals who were previously infected with SARS-CoV-2 seem to gain additional protection from a subsequent single-dose vaccine regimen. Though this finding corresponds to previous reports^24,25^, we could not demonstrate significance in our cohort.

The advantageous protection afforded by natural immunity that this analysis demonstrates could be explained by the more extensive immune response to the SARS-CoV-2 proteins than that generated by the anti-spike protein immune activation conferred by the vaccine^26,27^. However, as a correlate of protection is yet to be proven^1,28^, including the role of B-Cell^29^ and T-cell immunity^30,31^, this remains a hypothesis.

Our study has several limitations. First, as the Delta variant was the dominant strain in Israel during the outcome period, the decreased long-term protection of the vaccine compared to that afforded by previous infection cannot be ascertained against other strains. Second, our analysis addressed protection afforded solely by the BioNTech/Pfizer mRNA BNT162b2 vaccine, and therefore does not address other vaccines or long-term protection following a third dose, of which the deployment is underway in Israel. Additionally, as this is an observational real-world study, where PCR screening was not performed by protocol, we might be underestimating asymptomatic infections, as these individuals often do not get tested.

Lastly, although we controlled for age, sex, and region of residence, our results might be affected by differences between the groups in terms of health behaviors (such as social distancing and mask wearing), a possible confounder that was not assessed. As individuals with chronic illness were primarily vaccinated between December and February, confounding by indication needs to be considered; however, adjusting for obesity, cardiovascular disease, diabetes, hypertension, chronic kidney disease, chronic obstructive pulmonary disease, cancer and immunosuppression had only a small impact on the estimate of effect as compared to the unadjusted OR. Therefore, residual confounding by unmeasured factors is unlikely.

This analysis demonstrated that natural immunity affords longer lasting and stronger protection against infection, symptomatic disease and hospitalization due to the Delta variant of SARS-CoV-2, compared to the BNT162b2 two-dose vaccine-induced immunity. Notably, individuals who were previously infected with SARS-CoV-2 and given a single dose of the BNT162b2 vaccine gained additional protection against the Delta variant. The long-term protection provided by a third dose, recently administered in Israel, is still unknown.

## Tables and figures

**Table S1.** OR for COVID-19-related hospitalizations, model 1, previously infected vs. vaccinated

| Variable | Category | $\beta$ | OR<br>hospitalized | 95%CI | P-value |
| --- | --- | --- | --- | --- | --- |
| **Induced Immunity** |  |  |  |  |  |
|  | Previously infected | Ref |  |  |  |
|  | Vaccinated | 2.09 | 8.06 | 1.01 – 64.55 | 0.049 |
| **SES** |  | 0.05 | 1.05 | 0.72 – 1.53 | 0.81 |
| **Age $\geq 60$ yrs (16-39, ref)** |  | 5.08 | 160.9 | 19.91 – 1300.44 | <0.001 |

OR – Odds Ratio; SES – Socioeconomic status on a scale from 1 (lowest) to 10

**Table S2.** OR for COVID-19-related hospitalizations, model 2, previously infected vs. vaccinated

| Variable | Category | $\beta$ | OR<br>hospitalized | 95%CI | P-value |
| --- | --- | --- | --- | --- | --- |
| **Induced Immunity** |  |  |  |  |  |
|  | Previously infected | Ref |  |  |  |
|  | Vaccinated | 1.95 | 7.03 | 2.1 – 23.59 | 0.002 |
| **SES** |  | -0.07 | 0.93 | 0.74 – 1.17 | 0.547 |
| **Age $\geq 60$ yrs (16-39, ref)** |  | 4.3 | 73.5 | 25.09 – 215.29 | <0.001 |

OR – Odds Ratio; SES – Socioeconomic status on a scale from 1 (lowest) to 10

